# Efficiency of mobile eye camps for providing combined eye and vision care in underserved areas in Uttarakhand

**DOI:** 10.1101/2020.10.27.20217000

**Authors:** Siegfried Wahl, Alexander Leube, Renu Dhasmana, Premjeeth Moodbidri, Vasuki Krishna Kumar, Nitin Sisodia, Joachim Kuss

**Affiliations:** Institute for Ophthalmic Research, Eberhard Karls University, Elfriede-Aulhorn-Str. 7, Tuebingen 72076, Germany; Carl Zeiss Vision International GmbH, Turnstr. 27, Aalen 73430, Germany; Department of Ophthalmology, Himalayan Institute of Medical Science, Dehradun 248140, India; Carl Zeiss India Pvt. Ltd, Plot No.3, Jigani Link Road, Bangalore 560099, India; Sohum Innovation Lab, Whitefield, Bangalore 560066, India

**Keywords:** Eye care, Vision screening, India, Aloka Vision Program

## Abstract

To report eye examinations findings and cost-efficiency of mobile eye and vision care screening in underserved areas in north India. The Aloka Vision Program combines optometrical and ophthalmological screening as mobile eye camps with organized referrals to local eye hospitals. 402 people from urban (N = 191) and rural (N = 211) areas in the district of Uttarakhand, India, were screened for refractive error (RE), visual acuity (VA) and eye health. Statistical analysis was performed using ANOVA model and odd ratios. Costs were estimated based on the expenses of the camps. 44 % of the participants were male and 56 % were female and the age ranged from 7 to 72 years (urban) and 7 to 80 years (rural). Lack of accessibility of eye care was mentioned by 10% of the urban and 47% of the rural participants, why not attending regular vision test. Mild and severe visual impairment VA < 0.5 logMAR affected every fifth person, independent from the living environment. RE showed a myopic trend for the urban environment (Δ M = 0.67 D, p = 0.11). The risk for blindness was three times higher in rural compared to the urban area, mainly caused by cataract. The major costs are given by the treatments (58 %), followed by mobilization and organization (∼30 %), whereas personal costs are low (∼11%). Combined eye and vision care models reduce costs for separated screening’s organization and thus can increase the effectiveness of eye screening programs significantly.

## 1. Introduction

Globally, the leading causes of visual impairment are uncorrected refractive error (43 %)1 and cataract (33 %).[1] These two eye health related conditions are easily treatable by simple glasses or standard surgery resulting in at least 75 % of preventable cases of blindness.[2] Prevalence data for India from 2010 revealed that 133 million people, including 11 million children, are affected by visual impairment or blindness.[3] According to the World Health Organization (WHO), these are the second highest numbers in the world.1 Under the consideration of the WHO program “VISION 2020: The Right to Sight”,[4] focused screening programs can increase the awareness of visual impairment and blindness and further improve the availability of vision care service [5] to eliminate avoidable blindness by the year 2020.[4] Governmental programs in India adopted the goals of VISION 2020 aim in the twelfth five year plan to establish 5,000 vision centers for primary eye care by 2017 which serve as local first interface points.[6] Additionally, to these programs, initiatives from non-governmental organizations (NGOs) and the private sector can enhance the efficacy and reachability of eye care services.[6] Mobile eye care programs like the “Nayantara” project from Tirupati Eye Centre [7] uses traveling specialists to diagnose and treat eye conditions, e.g. diabetic retinopathy. Tele-Ophthalmology approaches as the Aravind Tele-Ophthalmology Network [8] or the Sankara Netralaya Tele-Ophthalmology Project [9] can further improve primary eye care with keeping patient satisfaction high, especially in rural areas.[10] Effectiveness and reliability of tele-ophthalmogy models were shown for the detection of diabetic retinopathy [11] but seem to be not appropriate for determination of refractive errors. Next to an increase in efficiency and availability of primary eye care services, there is a further need on trained and educated specialists which could provide this eye care services.[12]

The Aloka Vision Programme screening scheme, shown in Figure 1, combines a training of local entrepreneurs with the availability of primary eye care in providing ophthalmologic and optometric screening with an end-to-end support of dispensing spectacles or medical eye care. The focus is on an autonomous and self-energizing process to establish eye camps and vision correction for underprivileged people. The current study reports on frequency estimations of refractive errors and eye health conditions and compares cost-efficiency of a combined eye and vision care services in underserved areas in India.

**Figure 1:**
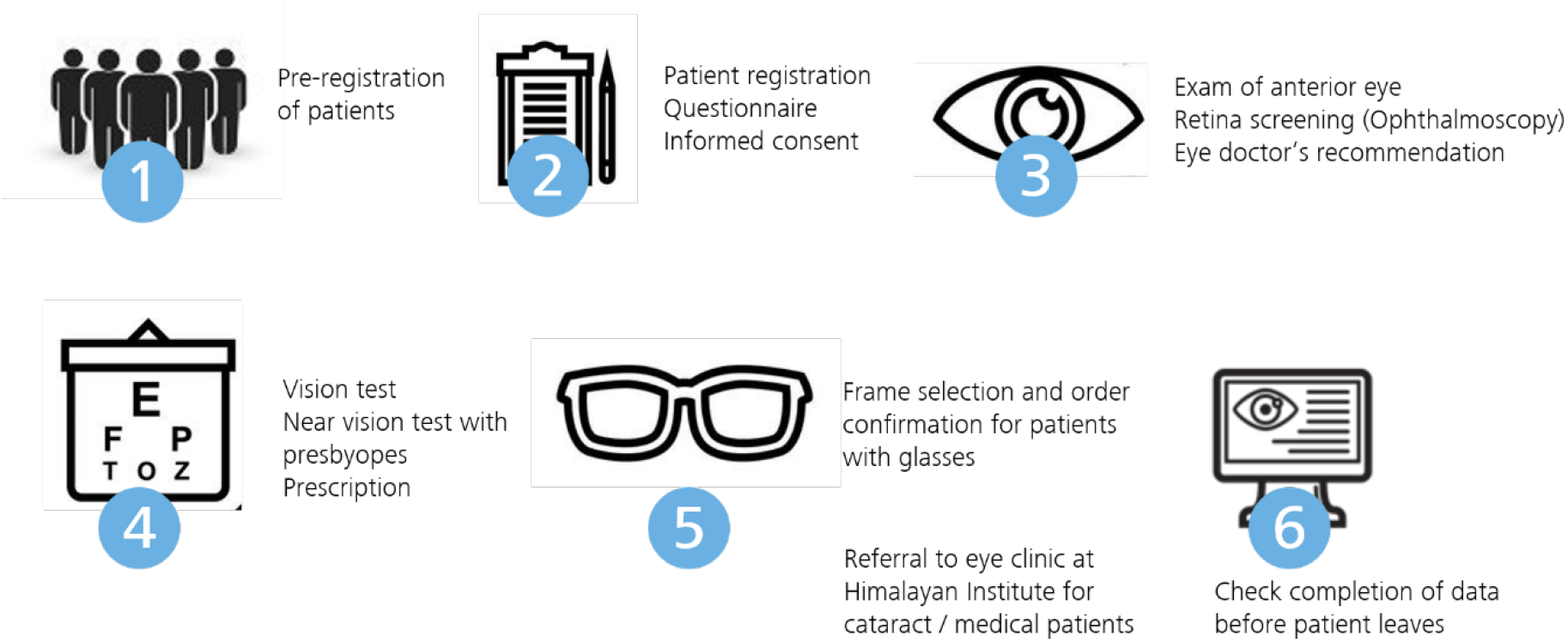
The Aloka Vision Programme screening scheme

## 2. Materials and Methods

### Participants and Public Involvement

The current study reports on vision screenings and their associated costs on behalf of the Aloka Vision Program carried out in June 2018 in the northern India State of Uttarakhand. Vision screening was performed by setting up mobile eye camps in an urban area (Dehradun city, 450 m above sea height) and in a rural area (Mussoorie, 2,000 m above sea height). Each eye camp included one to two optometrists from Carl Zeiss India Pvt. Ltd, one ophthalmologists from the Department of Ophthalmology, Himalayan Institute of Medical Science, volunteers from local NGOs and supporting stuff. The eye camps were open for everybody and free of charge. The study design follows prospective, cross-section, case-finding screening program.[13]

### Screening Protocol

Prior to the participation in the study, the screening protocol included an informed consent which was provided to the participants or in case of age under 18 by a parent or legal guardian. Further, the rationales of the study in accordance with the Declaration of Helsinki. Ethical considerations were ensured, since the Aloka Vision Program is part of the general eye camps performed by the eye doctors from the Himalayan Institute of Medical Sciences, Department of Ophthalmology. Study documents were in English language and explained in the local language. After registration participants were asked to fill a questionnaire on their educational level, their occupation, time spent outdoor, the status of spectacles correction, the awareness of vision care and the knowledge about healthy impact of ultraviolet radiation (UVR). Standardized protocol was followed by examinations from ophthalmologists in regards of anterior and posterior eye health. Possible eye conditions of pterygium [14] and cataract [15] were graded based on standard classification scales. Retinal fundus evaluation was performed using direct ophthalmoscopy and retinal abnormalities were graded by a trained ophthalmologist. After ophthalmological service, assessment of unaided visual acuity (VA), monocularly, using a printed eye chart with numbers or Tumbling E on a distance of 20 feet (6 m). Objective refraction for far vision was carried out using retinoscopy and was followed by a subjective refinement using a trial frame and trial lenses. Near vision test was performed in a distance of 40 cm to assess the additional power. For participants with uncorrected refractive error for far and/or near vision, a small selection of different optical frames was provided directly at the eye camp side. Later, glasses were produced by local opticians. In case of necessary surgery or further ophthalmological examination, participants were referred to the local ophthalmology clinic. Delivery of the glasses and transportation to the hospital was organized and carried out by local NGO partner.

### Statistical analysis

For statistical analysis, the results from subjective refraction were transformed from sphere, cylinder and axis notation to their power vector components. [16] Frequency data on the incidence of uncorrected refractive error was calculated for the mean spherical refractive error M and classified as myopic for M ≤ -0.5 D, as hyperopic for M ≥ +0.5 D and otherwise as emmetropic. Astigmatic refractive errors are considered as negative cylindrical errors Cyl ≤ - 0.5 D. Visual acuity is transformed to a logarithmic scale (logMAR) and classified according to WHO scale1 as normal vision for VA ≤ 0.0 logMAR, mild and severe visual impairment for 0.5 ≤ VA ≤ 1.3 logMAR and as blindness for VA > 1.3 logMAR. Analysis of eye health status and near vision needs were analyzed for participants aged >40 years (N = 270: Nurban = 134, Nrural = 136). To analyze differences between the urban and the rural area, analysis of variance (ANOVA) was conducted for refraction and visual acuity data using the location as parameter. In case of categorical grading of diseases severeness and data from the questionnaire, differences were analyzed using the χ^2^-test. The significance level was set to α = 0.05. To determine the risk suffering from visual impairment, refractive error or ocular health conditions between the two screening locations, odd ratios were calculated. [17] All calculations were done in a matrix-based program (Matlab 2018a, Mathworks).

## 3. Results

A total number of 402 people were screened, including 191 participants in the urban and 211 participants in the rural area. 44 % and 43 % were male and 56 % and 57 % were female for the urban and rural area, respectively. Since the participation in the eye camp was open for all age classes, the age of participants ranged from 7 to 72 years and 7 to 80 years in the urban and rural area, respectively. Age frequency distribution for both eye camps locations are shown in Figure 2.

**Figure 2:**
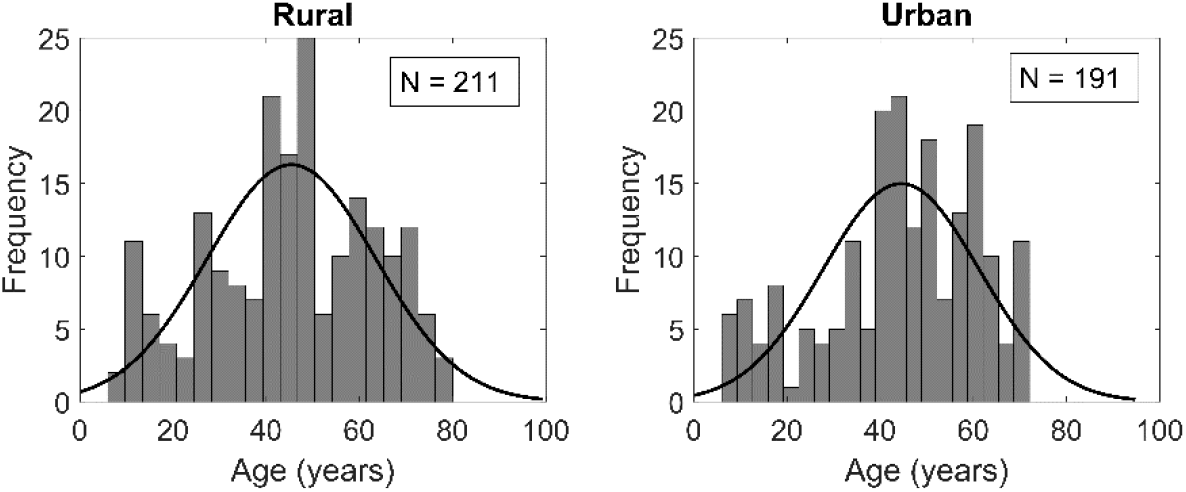
Age (years) distribution of eye camp locations in the urban (Dehradun) and rural (Mussoorie) area

### Eye healthy status

Additionally, to the optometric examinations, measuring refractive errors and visual acuity, the medical integrity of the participants eyes was checked by an ophthalmologist. As shown in Table 1, frequency for pterygium and cataract was analyzed as two abnormal structural changes of the anterior eye segment and general abnormalities from the posterior segment. Frequency of pterygium and cataract, calculated for the entire screened population does not show a significant difference between the rural and urban area (χ^2^ = 5.05, p = 0.17; χ^2^ = 3.89, p = 0.56). When analysis was performed only for participants aged older 40 years, no significant difference could be revealed (χ^2^ = 0.27, p = 0.61; χ^2^ = 4.66, p = 0.45). The main cause of blindness for the study population was cataract (n = 4, 80 %) followed by high myopia (n = 1, 20 %). Frequency of abnormalities from the fundus examination included conditions like exudates or drusen, vascular changes as retinal haemorrhage or glaucomatous changes of the optic disc. Living in a rural environment increased the risk of retinal abnormalities by 2.2 and 3.3 for the entire population and for the older population aged > 40 years, respectively.

**Table 1:** Frequency data on unaided visual acuity (VA), type of refractive error (D) specified by the spherical equivalent (M) and ocular health for rural (Mussoorie) and urban (Dehradun) screening locations in north India. Odd ratios (OR) are calculated between the two locations

|  |  | All n (%) |  | OR | Age >40 n (%) |  | OR |
| --- | --- | --- | --- | --- | --- | --- | --- |
|  |  | Rural | Urban |  | Rural | Urban |  |
| VA (N = 362) | Normal vision (VA ≤ 0.0) | 99 (53%) | 79 (44%) | 1.01 | 50 (40%) | 47 (39%) | 1.08 |
|  | MSVI (0.5 ≤ VA ≤ 1.3) | 40 (21%) | 38 (21%) | 0.93 | 37 (29%) | 30 (24%) | 1.31 |
|  | Blind (VA > 1.3) | 4 (2%) | 1 (0.6%) | 3.89 | 3 (2%) | 1 (0.8%) | 3.00 |
| Spherical equivalent (N = 402) | Hyperopia (M > 0.5 D) | 49 (23%) | 40 (21%) | 1.14 | 47 (35%) | 37 (28%) | 1.38 |
|  | Emmetropia (M ± 0.5 D) | 142 (67%) | 117 (61%) | 1.30 | 76 (56%) | 83 (62%) | 2.89 |
|  | Myopia (M < 0.5 D) | 20 (10%) | 34 (18%) | 2.07 | 13 (9%) | 14 (10%) | 1.31 |
| Astigmatism | Cylinder ≤ -0.5 D | 25 (12%) | 26 (14%) | 1.17 | 18 (13%) | 15 (11%) | 1.21 |
| Pterygium | OD | 13 (7%) | 6 (3%) | 2.02 | 8 (6%) | 9 (7%) | 1.15 |
|  | OS | 10 (5%) | 9 (5%) | 1.01 | 6 (5%) | 4 (3%) | 1.50 |
| Cataract (NOC > 1) | OD | 26 (14%) | 33 (19%) | 1.48 | 24 (19%) | 33 (27%) | 1.52 |
|  | OS | 20 (11%) | 32 (18%) | 1.92 | 19 (15%) | 32 (26%) | 1.93 |
| Retina | No abnormalities | 202 (96%) | 174 (91%) | 2.20 | 205 (97%) | 174 (91%) | 3.30 |
|  | Abnormalities | 9 (4%) | 17 (9%) |  | 6 (3%) | 17 (9%) |  |

### Refractive error status

Table 1 provides a summary of the frequency on the unaided visual acuity and the spherical equivalent refractive error for far vision. Worsening of unaided visual acuity was statistical significantly correlated with an increase in age (r = 0.43, p < 0.01), although the explained variance was low R2 = 0.18. The difference in the mean spherical refractive error analyzed between the urban and rural area showed a small shift towards more negative values for the urban area (Δ M = 0.26 D, p = 0.11), but did not reach significance. Similar, the astigmatism components J0 and J45 were not significantly different between the urban and rural areas (p_J0_ = 0.07, p_J45_ = 0.68). When analyzing only ammetropic citizens (N = 133) this shift towards more myopic refractive errors become more pronounced and clinically significant, but not statistically significant (Δ M = 0.67 D, p = 0.11). Following the measurements of the refractive errors of the eye for far and near vision, participants were asked to select an optical frame to be able to manufacture corrective glasses either for far, near or combined correction. In total, 63 % and 61 % of the people in rural and urban area were required to wear vision correction.

Since not every medial or refractive conditions needs a treatment, analysis was further conducted for the frequency of referrals. As shown in Figure 3, four categories were analyzed: Prescription of glasses, cataract surgery, general referral to an eye hospital and no referral necessary. There was no significant different found between the rural and urban area in both age groups (All: χ^2^ = 5.58, p = 0.13; Aged > 40: χ^2^ = 3.81, p = 0.28). In general, Figure 3 reveals that a screening program aimed to discover uncorrected refractive errors would account for around 60 % of the causes for visual impairment, whereas a pure ophthalmologic screening program would account for around 30 % of the cases. A combined screening program of ophthalmology and optometry increases the number of discovered cases of visual impairment and enables to discover most cases suffering from eye health and vision conditions.

**Figure 3:**
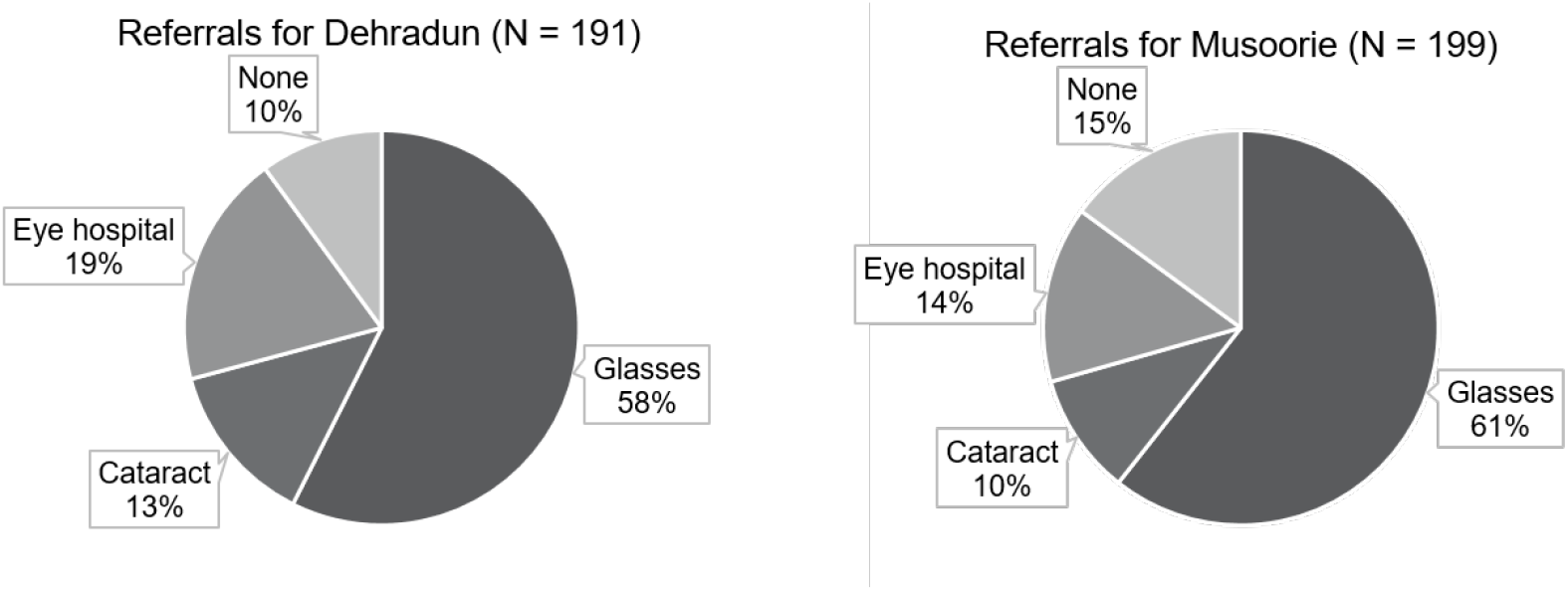
Frequency of different reasons for referral in the urban (Dehradun) and rural (Mussoorie) area

### Need for refractive and non-refractive treatment in aged sub-population

Presbyopia, the inability of near focusing of the crystalline lens, was analyzed for participants aged older 40 years (N_Urban_ = 134 and N_Rural_ = 136). 84 % of presbyopes required an additional power ≥ 1.0 D. Since there was no significant difference in age groups between the urban and rural area, no statistically significant difference could be found for the frequency and the amount of near addition power (Δ ADD = -0.17 D, p = 0.12).

The evaluation of the refractive error and the eye status for patients >40 years shows that, irrespective of pseudophakia or the possession of reading or distance glasses, only one of 247 was adequately supplied with near spectacles. This resulted in a frequency of 58 % in the rural and 61 % in the urban community which could be supplied with individually prescribed glasses. For 24 % in the rural and 32 % in the urban area, a medical treatment in the eye clinic followed by further prescription of glasses was arranged. For a group of 18 % respectively 7 % of the participants further dilated eye examination were required, the eye doctor’s and optometrist’s recommendation rejected by the person or as in case of retinal diseases no adequate medical treatment available.

With a combined eye care approach, a treatment of individual vision correction or sight preservation treatments has been arranged for 90 % while an optometry camp would have helped 60 % only, or an ophthalmological camp 28 % only.

### Cost estimations

Although the majority of the participants suffer from uncorrected refractive error or treatable ocular health conditions (see Table 1), analysis of the results from the questionnaire (multiple answers possible) revealed that 64 % in the urban area and 63 % in the rural area cannot see a benefit in regular vision tests or eye exams. In terms of accessibility and affordability, 10 % of people in the urban area further mention that vision care is not accessible, whereas in rural area accessibility was with 47 % the second main reason not attending regular eye tests. Only 25 % of people in the rural area were already wearing glasses, whereas the distribution of spectacles wear was with 36 % slightly higher in the urban environment. 7 % and 8 % of the people from the rural and urban area, respectively, underwent eye surgery before the screening for various reasons. According the national Reserve bank India, the poverty line for 2012 for the state Uttarakhand is 880 ₹ (∼12$) per month for the rural area and 1082 ₹ (∼14$) per month for the urban area. Poverty (% of people below poverty line) was shown to be significantly different (χ^2^ = 24.79, p < 0.001) between the urban (25 %) and the rural area (49 %), respectively. Whereas the educational level is not significantly different (χ^2^ = 3.41, p = 0.33).

The cost estimations for a four days inclusive eye camp are shown in Table 2. These costs represent the actual numbers from the reported camps and compromise next to the personal costs for the eye care experts and volunteers, the camp organization, the pre-promotion, people’s mobilization, camp support from local volunteers and further the costs for the respective treatment like spectacles or cataract surgery based on frequency values given in Figure 3. The major costs are given by the treatments (58 %), followed by mobilization and organization (∼30 %). The personal costs for the ophthalmologists and optometrist are low (11 %).

**Table 2:**
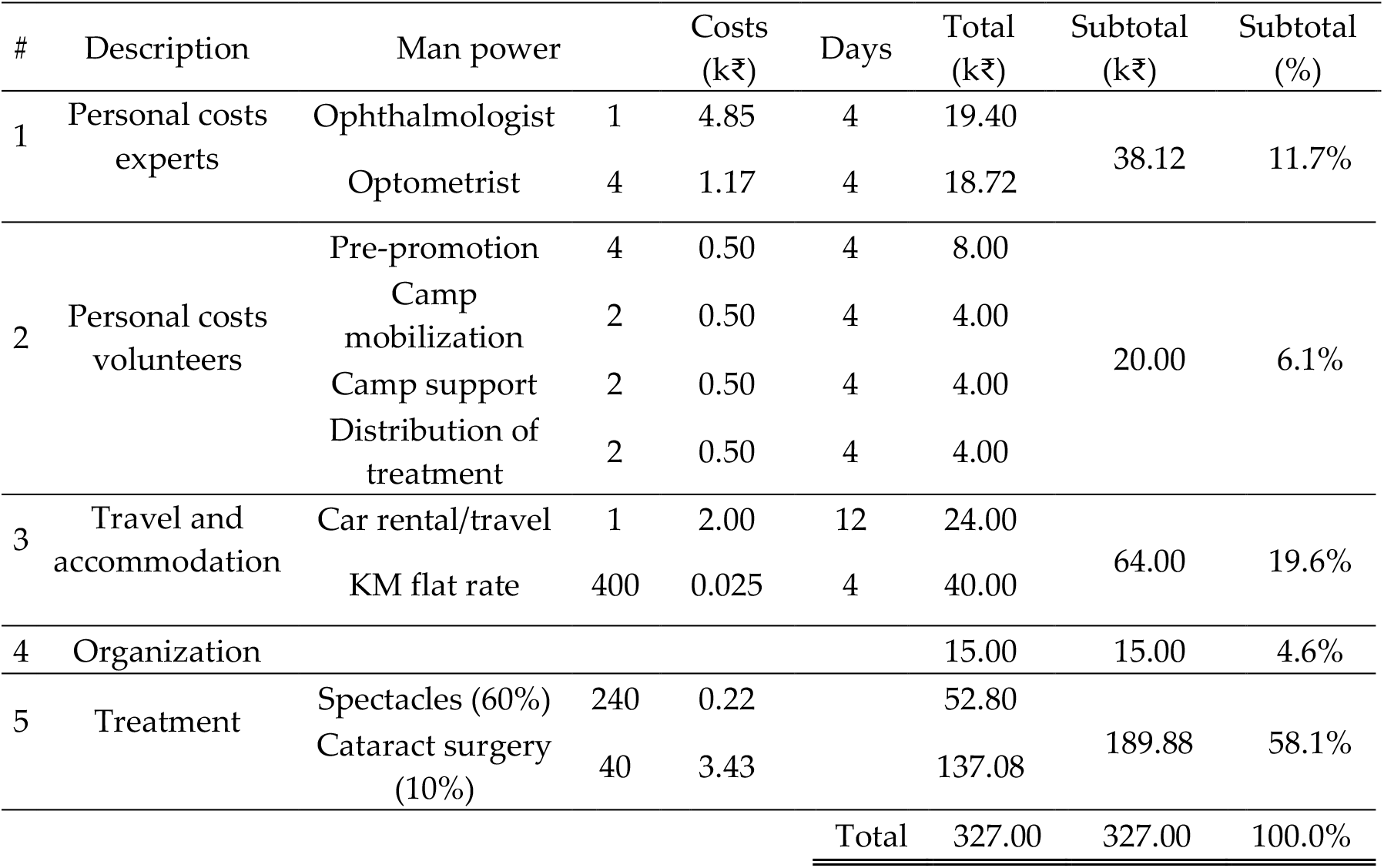
Cost estimation in k₹ for the pre-camp organization (4 days), completion (4 days) and treatment (4 days) of four days inclusive eye camps. The estimated numbers are based on the 400 screened people

| # | Description | Man power |  | Costs (k₹) | Days | Total (k₹) | Subtotal (k₹) | Subtotal (%) |
| --- | --- | --- | --- | --- | --- | --- | --- | --- |
| 1 | Personal costs experts | Ophthalmologist | 1 | 4.85 | 4 | 19.40 | 38.12 | 11.7% |
|  |  | Optometrist | 4 | 1.17 | 4 | 18.72 |  |  |
| 2 | Personal costs volunteers | Pre-promotion | 4 | 0.50 | 4 | 8.00 | 20.00 | 6.1% |
|  |  | Camp mobilization | 2 | 0.50 | 4 | 4.00 |  |  |
|  |  | Camp support | 2 | 0.50 | 4 | 4.00 |  |  |
|  |  | Distribution of treatment | 2 | 0.50 | 4 | 4.00 |  |  |
| 3 | Travel and accommodation | Car rental/travel | 1 | 2.00 | 12 | 24.00 | 64.00 | 19.6% |
|  |  | KM flat rate | 400 | 0.025 | 4 | 40.00 |  |  |
| 4 | Organization |  |  |  |  | 15.00 | 15.00 | 4.6% |
| 5 | Treatment | Spectacles (60%) | 240 | 0.22 |  | 52.80 | 189.88 | 58.1% |
|  |  | Cataract surgery (10%) | 40 | 3.43 |  | 137.08 |  |  |
| Total |  |  |  |  |  | 327.00 | 327.00 | 100.0% |

## 4. Discussion

The current study reports on a combined case-finding screening program in north India, Uttarakhand. Overall, the results reveal that a screening program aimed to discover uncorrected refractive errors would account for around 60 % of the causes for visual impairment, whereas a pure ophthalmologic screening program would account for around 30 % of the cases.

### Frequency of visual impairment and blindness

Visual impairment is classified by the WHO as visual acuity below 6/18 but above 3/60.[4] For visual acuity below 3/60 the person is classified as blind. Apart from the resolution performance of an eye, measured as the visual acuity, the functional visual field can be examined and is defined, in case of blindness as below 10°. The frequency reported from the current study on the visual impairment ∼ 20 % for rural and urban area in north India are higher as compared to the literature. The main reason for this is that the currently investigation is not a population-based study, since the no random cluster strategies are applied. However, Studies which were based on these random assignments found prevalence of 8 % [18], 10 % [19] and 2 % [20] of mild and server visual impairment among southern, western and north Indian populations, respectively. However, prevalence data can vary between different states, since a population-based survey from Murthy et al. (2005) in Gujarat, northwestern India, revealed a prevalence of visual impairment of 29 %.[5] The current screening program design results in a higher frequency of visual impairment findings due to the general openness of the program without any preliminary selection. Therefore, the likelihood for people without having subjective visual problems to join such screening programs is lower compared to people have visual complains. Dandona et al. (2002) conducted the Andhra Pradesh Eye Disease Study on 12,000 people from southern India and revealed that living in a rural area having a statistically significant higher odd as living in an urban area (odds ratio 2.12) to suffer from moderate visual impairment.[18] The current study revealed a higher odd only for the age group > 40 years, while this was associated with a higher risk for blindness when living in the rural area. The global trend of increased prevalence in urban environments of myopic refractive error, mainly studied in school children [21-23], can be confirmed in the current study (odds ratio 2.1), when analyzing the entire study population aged from 7 to 80 years.

Previous studies conducted rapid assessment of avoidable blindness (RAAB) for different countries in the world and revealed prevalence of blindness ranged from 1 % to 8 %.[24–27] Prevalence of blindness estimated from the current study for north India, showed comparable values and a higher risk for blindness in rural areas. due to the small sample size, the absolute numbers of blindness are low. The difference between the living environments is confirmed by previous findings from Patil et al. (2014).[19] There the prevalence of blindness was with 1.8 % significantly higher in rural areas in western India than in urban areas. This trend was shown also for other regions in the world like Indonesia [28], China [22] or Pakistan.[29] Globally, the main cause of blindness is cataract [1], as confirmed by the current study also for north India. Additionally, the age, higher life time exposure to UVR might be associated with an increased risk for cataract.[30–33] Furthermore, it is expectable that the combination of rural living environment, more time spent outdoors and higher altitude associated with higher UVR exposure [34] leads to an increase in UV-related eye disease like pterygium and senile cataract. However, odds from the current study showed only a slight increase of risk. A possible explanation is that UVR level can vary between urban and rural environments due to differences in the albedo between city surfaces and for instant grass or soil areas in rural areas. [35–38] A second reason for an elevated risk, specifically for cataract in the urban area, can be an economical one. Main reason for no eye care was affordability in the urban area. This is contradictory to the finding of BPL. However, the reported frequencies have to be interpreted cautiously since the current study design was an opportunistic screening and no cross-sectional population-based study.

### Socio-economic benefits of inclusivity of ophthalmology and optometry in vision screening programs

Assuming that the organizational parts in separated screening programs for eye health or vision care keeps similar, Table 2 shows that the travel expenses and costs for mobilization and coordination represent the largest share and therefore a combined eye and vision care program has a higher cost-efficiency.

The World Health Organization’s Vision 2020 program emphasizes to “strive to make refractive services and corrective spectacles affordable and available to the majority of the population through primary health care facilities, vision screening in schools and low-cost production of spectacles”.[4] Besides the fact that patients appreciate restoring of sight regardless ophthalmological treatment or providing glasses suits their needs, a cost efficiency ratio speaks for inclusion of eye and vision care as the major pool of costs (mobilization, local support, travel) can be spared by combining screening for refractive as for non-refractive errors and visual impairments.

Models for primary eye care in India utilize different aspects of vision screenings, covering for instance vision centers through permanent infrastructure [39,40] mobile units including screening and treatment abilities [7,9] or a combination of both.[6] One remarkable aspect of providing vision and eye care in unserved, rural and urban areas, is the patient satisfaction. In a review article Misra et al. (2015) pointed out that one important feature to satisfaction is the accessibility and the associated traveling costs.[6] Considering the fact that the next screening facility would require hour-long travel and, as the majority of workers in rural areas are day-laborers, the loss of a day income the expenses sum up to ₹850-₹1,000, compared to ₹220 for complete Aloka pair of spectacles including vision test.

The assessment of the refractive errors of an eye can lead to the treatment of the most prevalent cause for visual impairment.1 Data from the current study show that the spectacles coverage is higher in urban than in rural areas, which is in line with finding for other countries like Kenya [41], Bangladesh [42] or Australia.[43] This is mainly explained by the higher density of optical stores in cities. The finding from the current study, that there is no difference in the distribution of refractive error, but a lack of accessibility to corrective glasses, underlines the need of inclusive ophthalmological and optometrical vision care programs. Providing spectacles in unserved areas was shown previously to be a cost effectiveness intervention [44] for India [45] and other regions in the world.[46] Hereby, the costs of fitting and dispensing spectacle vary between studies and are estimated with US$5.0 [47] to US$25.0 [45], depending on the country. A RAAB from Marmamula, Keeffe and Rao (2009) in the district of Andhra Pradesh, India, revealed an unmet need for the correction of refractive errors for far and near vision.[48] The authors conclude that a delivery model like it provided by the vision centers [39,40] in conjunction with local optical outlets could serve as a successful vision care service. The training of local entrepreneurs within the scope of the Aloka Vision Programme and the results on the current frequency numbers on visual conditions enable a better effectiveness of spectacles coverage in urban and rural areas.

Cataract as leading cause of blindness worldwide1 represents a treatable eye health condition which deteriorates vision by the clouding of crystalline lens. Cataract surgical rate (CSR) was set by the Global Action Plan 2013 from the WHO as one core indicator for the access to cataract services in a country.[49] Wang et al. (2016) analyzed CSR from 152 countries and showed that there is a linear relationship between CSR and the gross domestic product per capita of a country.[50] The data for India shows a higher CSR as expected from the model and an increase in CSR from 4,800 to 6,000 in the years from 2009 to 2014 showing that the access to cataract service raised for the last years. Besides the availability of health care, these procedures have to be affordable by the people. For different regions in the world, cataract surgery was shown to be a cost-effective procedure to restore vision,51 but cost-Utility ($ / QALY, Dollar per quality adjusted life year) depends strongly on the individual country. In western countries like the United states, the cost-utility for cataract surgery is 2,020 US$ / QALY [51,52], whereas cost-utility for India is 8.07 US$ / QALY.[51,53] Comparing cost utility numbers with the willingness-to-pay showing that cataract surgery is an affordable treatment also in developing countries.

## 5. Conclusions

To conclude, the current study reports on a case-finding screening program as a combination of mobile examinations and hospital-based treatment referrals. Frequency of eye conditions showed that uncorrected refractive error account for most of visual impairments, which could be cost-effectively treated by spectacles fitting. A holistic approach providing combined ophthalmology and optometry examinations combined with a mobile screening camp and a well-organized referral coordination by local NGOs, private companies and local health partners, can provide an increase effectiveness for vision and eye care services in rural and urban areas in Uttarakhand.

## Supporting information

Aloka Ethics Approval Remote Eye Care

Aloka Ethics Proposal Remote Eye Care

## Data Availability

Data is available in the Ophthalmic Research Institute, University Tuebingen, Germany.

## Author Contributions

Conceptualization, S.W., A.L., R.D., P.M., N.S. and J.K.; methodology, S.W., A.L. and R.D.; formal analysis, S.W. and A.L.; investigation, S.W., A.L., R.D., P.M.,V.K.K., N.S. and J.K.; resources, S.W., R.D. and J.K.; data curation, A.L.; writing—original draft preparation, A.L.; writing—review and editing, S.W., A.L., R.D., P.M.,V.K.K., N.S. and J.K.; All authors have read and agreed to the published version of the manuscript.

## Acknowledgments

We would like to thank V. Kumar from Carl Zeiss India Pvt. Ltd, Nitin Sisodi from Sohum Innovation Lab, Parvitya Jan Kalyan Sansthan as non-governmental organization for the local organization and Petra Apelt from the Carl Zeiss Vision International GmbH for their continuous support. The ophthalmological examinations were performed by Dr. Shweta Sharma and Dr. Nishtha Yadav from the Department of Ophthalmology, Himalayan Institute of Medical Science. We further thank all participants for being part of this screening program.

