## Supplementary material for "Efficiency of mobile eye camps for providing combined eye and vision care in underserved areas in Uttarakhand": Aloka Ethics Approval Remote Eye Care

EMMANUEL HOSPITAL ASSOCIATION
808/92 DEEPALI BUILDING, NEHRU PLACE, NEW DELHI - 110019
FELLOWSHIP FOR TRANSFORMATION THROUGH CARING


### INSTITUTIONAL ETHICS COMMITTEE

#### Certificate of Approval

| | |
|---|---|
| Dr. Jacob Puliyel<br>*Chairman*<br><br>Mr. Siju Thomas<br>*Member (Legal)*<br><br>Ms. Sharmila Banerjee Livingston<br>*Member*<br><br>Dr. Oomen John<br>*Member*<br><br>Dr. Savita Doeme<br>*Member*<br><br>Dr. Jameela George<br>*Member Secretary* | **Title:** Remote eye screening study<br><br>**Researchers:** Dr. Vijayanand Ismavel, Dr. Roshine Mary Koshy, Ms. Ciin Hoih Kim, Mr. Joachim Kuss, Dr. Siegfried Wahl, Dr. Alexander Leube, Dr. Premjeeth Moodbidri<br><br>**Protocol No:** 209<br><br>**Comment:**<br><br>The study on "Remote eye screening study" was reviewed at the EHA IEC meeting on the 17th of May, 2019. The clarifications provided after the review have been accepted.<br><br>The protocol with clarifications given have been approved.<br><br>*The protocols are circulated to all members of the ethics committee prior to the scheduled meeting.* |

Approval is subject to the following conditions:

It is the principal investigators' responsibility to ensure that all researchers associated with this project are aware of the conditions of approval and which documents have been approved.

*All Serious adverse effects have to be informed to the* Secretary of the Institutional Ethics Committee*as per Good Clinical Practice (GCP) within 7 days. This approval stands automatically revoked if this is not complied with.*


Become an EHA Giving Partner - www.eha-health.org/give
CHARITABLE REGISTERED SOCIETY NO. 4546/1970-71 UNDER SOCIETY REGISTRATION ACT

Furthermore the principal investigator is required to notify the Secretary of the Institutional Ethics Committee the following:

1. Any significant change to the project and the reason for that change, including an indication of ethical implications (if any).
2. A delay of more than 12 months in the commencement of the project.
3. The inability of the principal researcher to continue in that role.
4. Any expiry of the insurance coverage provided with respect to sponsored clinical trials and proof of re-insurance;
5. Any unforeseen events or unexpected developments that could materially affect the approval
6. The Ethics Committee may conduct an audit/intermediate analysis if found necessary. All required data/documents must be provided to the Committee before further recruitment of cases;
7. Termination or closure of the project.

**Additionally, the principal researcher/investigator is required to submit**

- A Progress Report every 12 months for the duration of the project;
- A request for extension of the project prior to the expiry date, if applicable; and,
- A detailed Final Report at the conclusion of the project.

All research subject to the EHA Institutional Ethics Committee review must be conducted in accordance with the Ethical Guidelines for Biomedical Research on Human Participants published by Indian Council of Medical Research (ICMR).

**Special conditions**: None

Signed:.......Jameli George.......
30/5/14
Member Secretary, IEC

*(Please quote project no. and title in all correspondence)*
