## Supplementary material for "Efficiency of mobile eye camps for providing combined eye and vision care in underserved areas in Uttarakhand": Aloka Ethics Proposal Remote Eye Care

MAKUNDA CHRISTIAN LEPROSY & GENERAL HOSPITAL

Aloka Vision Programme

ZEISS

### Emmanuel Hospital Association

FELLOWSHIP FOR TRANSFORMATION THROUGH CARING

INSTITUTIONAL ETHICS COMMITTEE

APPLICATION FOR APPROVAL OF A RESEARCH PROJECT - 2019

Project Title: **Remote Eye Screening Study**

1. BASIC INFORMATION: Version No. 1

| | |
|---|---|
| Name of EHA Principal Investigator (Indian)<br>Address for correspondence:<br>Phone no:<br>E mail: | Dr Vijayanand Ismavel<br>Makunda Christian Leprosy and General Hospital<br>Bazaricherra 788727<br>Karimganj ,Assam<br><br>9365727334<br> |
| Other Principal Investigators:<br>E mail address:<br>Mobile No. | Dr Roshine Mary Koshy<br>Makunda Christian Leprosy and General Hospital<br>Bazaricherra 788727<br>Karimganj ,Assam<br><br>6000323503<br><br><br>Ms Ciin Hoih Kim<br>Department of Optometry<br>Makunda Christian Leprosy and General Hospital<br>Bazaricherra 788727<br>Karimganj ,Assam<br><br>9127259824<br> |
| Other Principal Investigators:<br>E mail address:<br>Mobile No. | 1. Joachim Kuss,<br>Head of Communications, ZEISS Consumer Markets Segment,ZEISS Vision Care<br><br>Carl Zeiss Vision International GmbH<br>Turnstrasse 27<br>73430 Aalen, Germany |

| | |
|---|---|
| | 2. Professor Dr Siegfried Wahl<br>Head of Advanced Development and Director ZEISS Vision Science Lab<br><br>ZEISS Vision Care<br>Carl Zeiss Vision International GmbH<br>Elfriede-Aulhorn-Straße 7<br>72076 Tuebingen, German<br><br>3. Dr Alexander Leube<br>Physiological Optics and Vision Scientist, ZEISS Vision Science Lab<br><br>ZEISS Vision Care<br>Carl Zeiss Vision International GmbH<br>Turnstrasse 27<br>73430 Aalen, Germany<br><br>4. Dr Premjeeth Moodbidri<br>Program Manager – Aloka<br><br>Carl Zeiss India (Bangalore) Pvt. Ltd.<br>ZEISS Group<br>Plot No 3, Jigani Link Road<br>Bommasandra Industrial Area<br>Bangalore – 560099, India |
| Collaborator:<br>Overseas | Zeiss Vision Science Lab |
| University: | University of Tubingen |
| Funding Agency / Sponsoring Organization:<br>Address:<br>Phone:<br>E mail: | Carl Zeiss India (Bangalore) Pvt. Ltd.<br>ZEISS Group<br>Plot No 3, Jigani Link Road<br>Bommasandra Industrial Area<br>Bangalore - 560099, India<br>Phone: +91 80 4343 8160<br>Mobile: +91 70228 58114 |
| Site contact details: [place(s) where research will take place]<br>Name:<br>Address:<br>Phone no:<br>E mail: | Department of Optometry<br>Makunda Christian Leprosy and General Hospital<br>Bazaricherra 788727<br>Karimganj ,Assam<br> |

| | |
|---|---|
| Approved budget for the research | Zeiss will bear the entire expense of the project |
| Research start date<br>Period of data collection<br>Research completion date | From June 1st 2019 to July 31st 2019 |
| Date of submission: | 9th May 2019 |

2. BACKGROUND INFORMATION:

Rationale for undertaking the research in the light of existing knowledge- (100-200 words)

Like in other Rapidly Developing Economies there is a lack of eye and vision care in rural areas of India which needs to be addressed. The Delhi Declaration 2010 states "it is estimated that almost half the population of India have some form of refractive error, and that around 133 million of them are blind or vision impaired as a result of lack of correction, including 11 million children".

The root cause for this shortage of regular and quality eye and vision care is the lack of trained optometrists and eye care professionals outside of urban areas.

With the Aloka Vision Program, ZEISS , a German manufacturer of optical systems and industrial measurement and medical devices, started a social business in 2015 to make affordable and quality eye and vision care available in unserved rural regions across India. In partnership with hospitals, non-governmental organizations and local entrepreneurs the Aloka team serves up to 8,000 patients per months, providing up to 3,000 glasses per month.

Since end 2018 the Aloka Vision Program has been collaborating with Makunda Christian Leprosy and General Hospital. Together the partners provide the hospital's patients with eye exams, vision tests and eyeglasses.

While this service enables available, affordable and quality vision correction, the partners aim to evaluate and test the expansion and further development of eye and vision care available at the hospital for the communities it serves. A promising way to do this can be so-called remote eye screenings, i.e. telemedicine technology for better eye and vision care.

3. AIMS/ OBJECTIVES/ OUTCOMES:

3.1 Hypothesis or research question:

3.2 Overall aim:

To evaluate the functionality, connectivity and satisfaction of patients with tele-medicine based eye examination and refraction services.

3.3 Objectives:

To determine if effective ophthalmic diagnosis is possible in remote rural locations in the absence of qualified on site optometrists / ophthalmologists using remotely controlled digital diagnostic equipment and tele ophthalmology

3.4 Expected Outcomes:

To show that convenient, safe, effective and affordable ophthalmic diagnostic services are possible through tele ophthalmology.

4. STUDY DESIGN:

Give a detailed description of the methodology of the proposed research (1-2 pages).

The methodology adopted will be a pilot phase cross sectional observational study conducted in the optometry department of Makunda Christian Leprosy and General Hospital(MCLGH)

Time period: 1st June 2019 to 31st July 2019

The Aloka optometrist at the Makunda Christian Leprosy and General Hospital would be equipped with state-of-the-art examination and test devices which are connected via Internet to remote ophthalmological and optometry experts for full eye exam and comprehensive vision test.

The installed platform at the hospital will include Slit Lamp for examination of the anterior eye ( ZEISS SL), fundus camera from ZEISS for examination of the posterior eye/retina (ZEISS Visuscout), Autorefractor for objective monocular refraction( ZEISS Visuref) and Phoropter with screen for subjective refraction (ZEISS Visuphor with Visuplan).

The remote team will consist of fully professionally trained optometrist of the Aloka Vision Program at ZEISS India, Bengaluru, Karnataka and the remote retina screening will include online services provided by ZEISS India.

The work flow is depicted as follows:

MAKUNDA CHRISTIAN LEPROSY & GENERAL HOSPITAL

Aloka Vision Programme

ZEISS

#### Remote Eye Screening Study at Makunda Hospital
#### Screening process

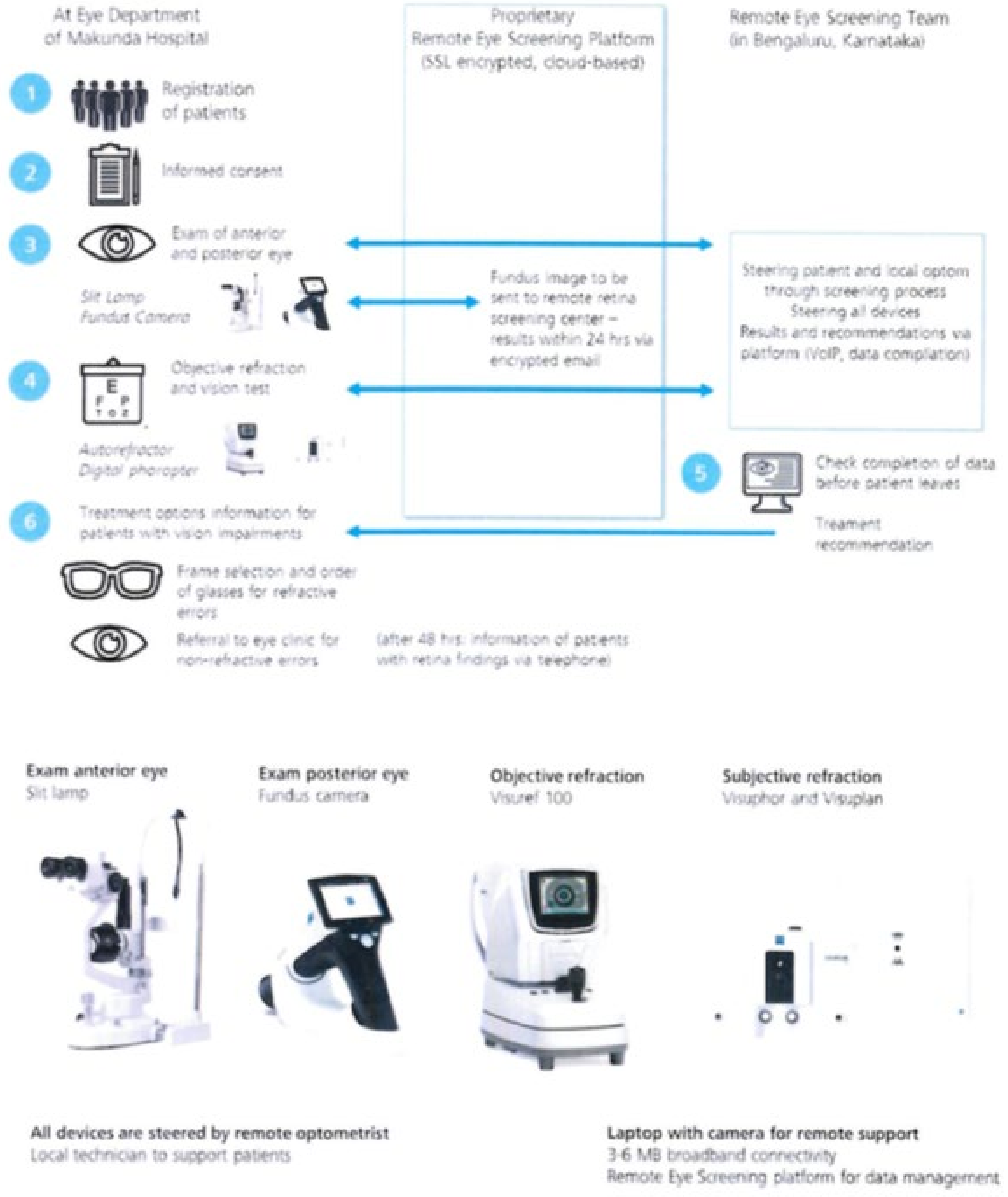

4.1 Participant recruitment procedure: Who are the research participants? (Eg: adult men, adolescents, age etc.). Explain how the participants will be recruited.

The research participants include all patients above the age of 8 years who have been referred by a Medical Officer from the out patient and in patient department of MCLGH to the Optometry department.

4.2 What are the Inclusion and exclusion criteria for entry of participants in the research?

Inclusion criteria:

All patients above the age of 8 years who are able to sit still and obey instructions given by the optometrist.

Exclusion criteria:

1. Patients below the age of 8 years
2. Patients who have not consented to the study

4.3 Inclusion and exclusion criteria for control participants, if any:

Not applicable

4.4 Number of Participants:

a. Sample size for quantitative date collection: What is the sample size? Justify the sample size. What sample size calculations were done to determine the number of participants? Mention the plans for statistical analysis of the study.

Observational study with a sample size of 1550.

All patients who are seen in the optometry department during the period of the study and who satisfy the inclusion and exclusion criteria are included as participants.

b. Qualitative data collection:

Self-administered questionnaire: Attached as annexure

5. Risks and Benefits:

5.1. What are the potential risks to the participants? Consider social and emotional risks as well as more obvious physical risks.

No risks involved

5.2. What is your risk management plan?

Not applicable

5.3. What will be the compensation for unexpected risks?

Not applicable

5.4. What are the potential benefits to the participants?

Patients will obtain high quality tele ophthalmology services, both for diagnostic and therapeutic recommendations from a team of ophthalmologist and professionally trained optometrists of the Aloka Vision program at Zeiss India, Bengaluru, Karnataka.

All patients where needed and within the range of available treatments will be offered a treatment based on the recommendation as result of the eye exam and vision test – individual and quality eye glasses for refractive errors or referral to eye clinic for further medical evaluation or eye surgery.

6. SAFETY AND OTHER CONTROLS:

6.1 Does this study involve ionizing radiation, hazardous substances, or hazardous or invasive procedures (including radiological imaging, vein puncture, or intimate physical examination)?

No
If yes, please justify:

MAKUNDA CHRISTIAN LEPROSY & GENERAL HOSPITAL

Aloka Vision Programme

ZEISS

7. INFORMED CONSENT:

| |
|---|
| 7.1 What will be the procedure for seeking Informed Consent from research participants? How will "research", "randomization", "risks and benefits" be explained?<br><br>The patient will be provided a Patient Information sheet which describes the details of the study. The contact details of the Principal Investigator will be also be provided to the participant for further clarifications.<br><br>7.2 What will be the procedure for seeking Informed Consent from parents/guardians etc of research participants who are children, mentally/ physically challenged? How will assent be obtained from these research participants?<br><br>For children less than 12 years, the patient's guardian will be provided a Patient Information sheet which describes the details of the study. The contact details of the Principal Investigator will be also be provided to the participant for further clarifications.<br><br>7.3. How will it be made clear that participants are under no compulsion to participate and may withdraw at any time without jeopardizing any service delivery or their relationship with the researcher?<br><br>It has been detailed in the patient information sheet that participation in the project is completely voluntary and participants can withdraw from the study at any point in time without affecting the evaluation and management of their medical complaint or their professional rapport with the treating physician.<br><br>7.4. Details of consent if the participants are audio –taped / video-taped?<br>Not applicable<br>7.5. Provide a copy of Plain Language Statement and Informed Consent Forms in English.<br>Attached as annexures<br><br>7.6. Details of proposed compensation and reimbursement of incidental expenses.<br>Not applicable<br><br>7.7. Statement of probable ethical issues and steps taken to tackle the same.<br>No ethical issues |

8. CONFIDENTIALITY:

| |
|---|
| 8.1 What procedures will ensure the confidentiality of participants?<br><br>All data that is being used for the project will be anonymized. |

8.2 The raw data collected will be locked and protected. I agree
8.3 The electronic data will be pass -word protected. I agree

9. OWNERSHIP & STORAGE OF DATA:

9.1 The data collected during the research will be stored and maintained by EHA Principal Investigator. The other Principal Investigators will have a copy of the data.

I agree

9.2 All the Principal Investigators will be responsible for the safety of the data.

I agree

10. POTENTIAL CONFLICT OF INTEREST: (ANY FINANCIAL INTEREST FOR RESEARCHERS)

Conflict of Interest: No

11. REPORT:

11.1 The research report will be finalized after all the principal investigators agree with the report.
I agree

12. PLANS FOR PUBLICATION & DISSEMINATION:

12. 1 All the Principal Investigators with mutual agreement will publish Articles/ Reports.
I agree

12. 2 How will results be disseminated? What information will be fed back to the participants and/or participating organization?
The result of the study will be used by partners in mutual agreement for scientific paper (incl. presentation at conferences and publication in peer-reviewed: magazine), for internal

| |
|---|
| and external communication. All communication materials will be approved by the hospital 'and by ZEISS before release. |

13. Annexures:

a. Patient information sheet
b. Consent form
c. Case record file
d. Recent CV of Investigators indicating qualification & experience particularly in research

**Annexure a**

#### Patient Information Sheet

MAKUNDA CHRISTIAN LEPROSY & GENERAL HOSPITAL | Aloka Vision Programme | ZEISS

Dear Sir or Madam,

I am representing the Aloka Vision Programme from Carl Zeiss India (Bangalore) Pvt Ltd. We are conducting an assessment of the Remote Eye Screening service developed by ZEISS. We would require some information about yourself, your medical history and awareness about eye check-up. We will protect this information from any unauthorized disclosure, tampering, or damage. We will be using this information to evaluate the efficiency and convenience of the Remote Eye Screening. All your personal data (prescription, name and delivery address) will only be used to dispense your spectacles if needed or for referral purposes. Your data will be used for scientific study and statistical purposes only in anonymised way without reporting any personal data. We will neither publish your personal data nor share it with third parties.

Refraction is the procedure in which we determine the best corrected optical lens of each eye for purposes of medical evaluation or for prescribing glasses. Refraction is necessary to adequately determine visual function as for instance acuity and is important in making sure that serious underlying eye problems do not exist. We perform refractions as a part of all our comprehensive eye evaluations, which are remotely controlled by certified eye doctors and optometrists. You participation in this study is completely voluntary and you can withdraw you participation agreement at any time without having any disadvantages

You will be provided with affordable quality spectacles if deemed necessary. If any other treatable diseases are diagnosed, we will refer you to an identified hospital which might require surgery or any other procedure. This service also may be charged depending on the hospital policies. If request by you, we will provide you with the results of the eye examination.

You can contact the principal investigator for further clarifications : Dr Roshine Mary Koshy / Phone no: 7035388537

Patient Acknowledgement

I have read the above information and understand that spectacles and any treatment may be paid. I accept full responsibility for the cost of further treatment and understand payment is due at the time of service if requested.

MAKUNDA CHRISTIAN LEPROSY & GENERAL HOSPITAL

Aloka Vision Programme

ZEISS

**Annexure b**

#### Consent form for Refraction and Treatment

I hereby voluntarily consent to the rendering of care by the doctors and authorized members of Carl Zeiss India (Bangalore) Pvt Ltd and Makunda Christian Leprosy and General Hospital, Bazaricherra including diagnostic procedures and medical treatment as necessary in their professional judgment.

I hereby acknowledge that no guarantees have been made to me as to the effect of such examinations or treatment to my condition.

I acknowledge that I am responsible for all reasonable charges in connection with care and treatment which are not part of the Remote Eye Examination, I attend, and which are provided by partners.

I have read this form and certify that I understand it contents.

Signature or Thumb Impression and name of participant/ guardian ( include nature of relationship )

________________________________________________

Signature and name of Witness (In case of illiterate patient):

Date :

Place: Makunda

MAKUNDA CHRISTIAN LEPROSY & GENERAL HOSPITAL

Aloka Vision Programme

ZEISS

Annexure c

### Case Record File
#### Personal information

| | | | |
|---|---|---|---|
| 1. | **Hospital Record Number** | | |
| 2. | **Age (in Completed years)** | | |
| 3. | **Gender** | ☐ M | ☐ F |
| 4. | **Education** | ☐ Did not go to school | ☐ Some formal schooling |
| | | ☐ SSC/HSC/10th Standard | ☐ Graduation |
| 5. | **Occupation** | ☐ Farmer | ☐ Driver |
| | | ☐ Labourer | ☐ Office work |
| | | ☐ Housewife | ☐ Others, please specify: |
| 6. | **BPL Status** | ☐ Yes | ☐ No |
| 7. | Main **complaints** regarding sight and eye - please list | 1. ____________<br>2. ____________<br>3. ____________ | |
| 8. | **Ocular history** (Any surgeries undergone) | ☐ Yes | ☐ No |
| 9. | **How often do you undergo eye check-up?** | ☐ Every year | ☐ Rarely |
| | | ☐ Every two years | ☐ Never |
| 10. | **If less than once in 2 years: What prevents you from doing eye check-up frequently?** | ☐ I am not convinced it is necessary / important for me<br>☐ Next eye check facility too far away / not accessible<br>☐ Costs of eye check to high / unknown | |

MAKUNDA
CHRISTIAN LEPROSY &
GENERAL HOSPITAL

Aloka
Vision Programme

ZEISS

**11. How comfortable you were with the vision screening experience?**

☐ I was very comfortable
☐ Comfortable
☐ Neither comfortable nor uncomfortable
☐ Uncomfortable
☐ Very Uncomfortable
☐ If uncomfortable please specify why

----------------------------------------------------------------

----------------------------------------------------------------

----------------------------------------------------------------

**12. Would you recommend remote eye screening to your familiy**

☐ Yes
☐ No
☐ Don't know

**13. How much would you be willing or able to spend for a full eye exam and vision test?**

☐ Nothing
☐ 20-40 INR
☐ 50-90 INR
☐ 100-140 INR
☐ 150-200 INR
